# Prevalence of Depression and Anxiety and Their Sociodemographic, Stigma, and Help-Seeking Correlates Among Adults in Bangladesh: A Multi-Site Cross-Sectional Study

**DOI:** 10.64898/2026.09.12.26362706

**Authors:** Shabbir Abdullah Maruf, MD Habibur Rahman, Most Sakila Azmin, Tasfia Afrin Faria, Md. Al Mamunur Rashid, Md Masum Billah, Nishat Subaha Siddique, Mohammad Biyazid Khan

**Affiliations:** Department of Sociology, Bangladesh University of Professionals (BUP); Department of Psychology, Teesside University, Middlesbrough, UK; Lecturer, Department of Business Administration, Uttara University; Department of Sociology, Noakhali Science and Technology University (NSTU); Institute of Social Welfare and Research, University of Dhaka; National University, Bangladesh; Lecturer, Green University of Bangladesh

**Keywords:** Depression, anxiety, stigma, help-seeking attitudes, Bangladesh, prevalence, cross-sectional study, psychosocial determinants, mental health, low- and middle-income countries

## Abstract

**Background:** There is a significant health burden of depression and anxiety in Bangladesh, but there is limited data on depression and anxiety prevalence in the community using multi-site studies that include psychosocial determinants. The independent association of mental health stigma and help-seeking attitudes with risk should be explored.

**Objectives:** To estimate the prevalence of moderate-to-severe depression and anxiety among adults in Bangladesh and examine their associations with sociodemographic characteristics, mental health stigma, and attitudes toward seeking professional psychological help.

**Methods:** A cross-sectional study was conducted from February 1 to June 20, 2026, across three divisions of Bangladesh (urban Dhaka, semi-urban Chattogram, rural Sylhet). Stratified multistage sampling recruited 400 adults aged 18–60 years from government primary care facilities. Depression and anxiety were assessed using the Patient Health Questionnaire-9 (PHQ-9) and Generalized Anxiety Disorder-7 (GAD-7), respectively (cutoff ≥10 for moderate-to-severe). Mental health stigma was measured using the Community Attitudes Toward Mental Illness—Bangla (CAMI-B), and help-seeking attitudes using the Attitudes Toward Seeking Professional Psychological Help—Short Form (ATSPPH-SF). Multivariable binary logistic regression examined independent predictors of depression and anxiety, controlling for sociodemographic variables.

**Results:** Depression prevalence was 39.0% (n = 156; M = 8.13, SD = 5.72) and anxiety prevalence was 29.8% (n = 119; M = 7.21, SD = 5.14). In adjusted models, each additional unit of CAMI-B stigma increased odds of depression (adjusted odds ratio [aOR] = 1.08, 95% confidence interval [CI] [1.03–1.13], p = .001) and anxiety (aOR = 1.07, 95% CI [1.02–1.12], p = .004). Each additional unit of positive help-seeking attitude reduced the odds of depression (aOR = 0.92, 95% CI [0.88–0.97], p = .001) and anxiety (aOR = 0.93, 95% CI [0.89–0.97], p = .001). Age predicted depression (aOR = 1.04 per year, p = .009) but not anxiety (aOR = 1.02, p = .439). Sex, education, and household income were not significant independent predictors in adjusted models. Descriptive analysis showed higher depression prevalence in rural (47.0%) versus urban (33.8%) settings, though rural residence was not significant in adjusted models.

**Conclusions:** Depression and anxiety are highly prevalent among Bangladeshi adults. Mental health stigma and positive help-seeking attitudes emerge as independent psychological determinants, stronger than conventional sociodemographic factors. These modifiable psychosocial factors represent key intervention targets. Comprehensive strategies combining community-level stigma reduction, individual help-seeking enhancement, and healthcare system strengthening are essential to reduce depression and anxiety burden and address Bangladesh’s substantial mental health treatment gap.

## 1. Introduction

Depression and anxiety are among the most common mental disorders worldwide and are important causes of disease burden (GBD 2019 Mental Disorders Collaborators, 2022; Wang et al., 2025). Most of them appear in late adolescence and early adulthood (around age 15) and are usually observed by this age (Chodavadia et al., 2023). They markedly disrupt quality of life, increase disability, and cause an estimated loss of productivity globally of US$1 trillion every year (Chodavadia et al., 2023; Jain et al., 2022). The burden was further increased by the COVID-19 pandemic, as estimates of depression and anxiety went up to 27.6% and 25.6%, respectively (Santomauro et al., 2021; Zhang et al., 2025), making these disorders prominent public health priorities.

There is a significant mental health burden in Bangladesh. Based on the criteria of the DSM-5, psychiatric disorder prevalence was found to be 13.7% for adults in a national community survey, with estimates suggesting 18–19% of adults had mental disorders in 2019 (Alam et al., 2019; Hasan et al., 2021; Suanrueang & Peltzer, 2025). The prevalence of depression was estimated at 5.2% – 6.7% using a diagnostic approach and 16.3% using a screening approach with a diagnostic approach, and the prevalence of anxiety was 3.5%– 4.7% and 6.0% using a screening approach, respectively (Pengpid et al., 2025; Sarkar et al., 2025, 2026). Adolescents, university students, married women, and older adults are among those who tend to have high depressive and anxiety symptoms (Habib et al., 2025; Haque et al., 2025; Islam et al., 2020, 2021; Rahman et al., 2025). Even with this burden, Bangladesh has a significant treatment gap, as only 3.9–4.1% of adults with mental disorders seek professional treatment and 92.3% do not receive treatment (Pengpid et al., 2025; Sarkar et al., 2026). Approximately 44.8% of affected women remain undiagnosed (Pengpid et al., 2025). Stigma, lack of services, limited funding, and mental health staffing (only 1.1 mental health professionals per 100,000 population) are key barriers (Haque et al., 2025; Pengpid et al., 2025).

Overall, there is a lack of discussion around mental illness in Bangladesh, and the general public has a strong stigma towards it, misconceptions about the cause, and a sense of social distance that can hinder help-seeking, and is associated with increased levels of depression and anxiety (Faruk et al., 2023; Roy & Chowdhury, 2024). The stigmatized attitudes are more prevalent among the urban residents, perhaps because of their increased social isolation and contact with mental illness (Faruk et al., 2023). There is social stigma associated with anxiety, depression and stress in discussion and these disorders are also under diagnosed clinically (Rahman, Biswas et all, 2025). These are additional challenges for women, such as limited health autonomy, fear of being labelled “rebellious,” and concerns about marital stability, which all deter women from seeking help and exacerbate their symptoms (Raza et al., 2025; Walter et al., 2025). A positive relationship exists between community attitudes towards mental illness and psychological distress across cultures (Alluhaibi & Awadalla, 2022).

The awareness of mental health issues is extremely low in Bangladesh, with only 8.5% understanding depression and 6.2% anxiety. (Uddin et al., 2019) It may lead to lack of awareness and delayed identification and help-seeking, which can foster poor attitudes to seeking professional psychological help as symptoms progress. Notably, of those who are aware, almost all are in a positive attitude towards medical or psychological counseling (Uddin et al., 2019). However, in reality, people do not seek professional help often (Rahman, Biswas, et al., 2025). Early signs of mental illness can be overlooked by individuals in Bangladesh and they may not seek professional help when they are suffering from mental illness due to personal beliefs, cultural norms and assumptions about mental illness and stigma (Ahmed et al., 2025). The patterns highlight the need to take into account the prevalence of depression as well as anxiety, and attitudes towards professional psychological support services among Bangladeshi adults.

These findings are supported by some previous studies in Bangladesh that anxiety and depression are not uniformly distributed among sociodemographic groups (Amin et al., 2025; Pengpid et al., 2025; Sarkar et al., 2025; Hasan et al., 2021). There are consistent associations between sex and depression and anxiety, with females generally reporting higher levels of depression and anxiety than males, although some studies report no difference (Islam et al., 2020; Pavel et al., 2026; Rahman et al., 2025). Age relationship is different among the population, older age group related to higher burden of anxiety and depression in reproductive age female and older adults, while few studies have found a higher burden at younger age or mid-adolescent age among students and adolescents (Amin et al., 2025; Islam et al., 2020; Rahman et al., 2025). In a recent study conducted among the late adolescents (15-21 years) revealed that the depression and anxiety rates were found to be high among the males age group 19-21 (Khatun et al., 2025). Low education, parental education, low to moderate income and lack of family savings also have consistent links with poor mental health; low education, parental education, low to moderate income and absence of family savings link with higher depression and anxiety (Amin et al., 2025; Khatun et al., 2025). But one national study of these disorders did not show that they are restricted to the poorer or less educated (Amin et al., 2025). Marital disruption, bereavement, separation, widowed status and urban or semi-urban residence are correlates of marital disruption and residence while residence and symptoms are associated with widowed status, urban or semi-urban residence, and bereavement (Amin et al., 2025; Islam, 2019). Employment status and religious affiliation have also been linked with depression and anxiety, but the size and direction of relationship have been different in various populations (Habib et al., 2025; Rahman et al., 2025). Also, the depression and anxiety rate is higher in later adolescent children who are unmarried, rural, and lack formal education (Khatun et al., 2025). Bangladeshi results are consistent with those of other South Asian studies, which identified female sex, lower education status, lower socioeconomic status, marital disruption, chronic illness and social and urban stressors as common factors contributing to depression and anxiety; however, the findings were somewhat inconsistent (Arvind et al., 2019; Vidyasagaran et al., 2023).

Although there is increasing evidence, there is limited multi-site data on depression, anxiety and sociodemographic and stigma and help-seeking correlates among Bangladeshi adults. Previous research has been conducted on any specific group, which reduces the generalizability to the wider adult population (Habib et al., 2025; Islam et al., 2021; Pavel et al., 2026). Furthermore, numerous studies were undertaken at single districts, cities, campuses or occupational sites (e.g. in schools of Dhaka, Mymensingh, Savar, Jahangirnagar University, or one rural district) (Islam et al., 2021; Khatun et al., 2025; Pavel et al., 2026). Some studies found that the findings of the key sociodemographic factors such as age, education level, wealth and residence are not consistent (Amin et al., 2025; Suanrueang & Peltzer, 2025). While Bangladesh Demographic and Health Survey (BDHS) studies have enhanced depression and anxiety prevalence estimates, relatively few community-based multi-site studies have been conducted that used validated instruments for depression (PHQ-9) and anxiety (GAD-7) and measured community attitudes towards mental illness and help-seeking attitudes (Amin et al., 2025; Esteban-Gonzalo et al., 2026; Rahman et al., 2025). In addition, there are no multi-site studies including urban, semi-urban and rural areas, which would help to understand regional differences in depression and anxiety.

The purpose of this multi-site cross-sectional study was to estimate the prevalence of depression and anxiety among adults in Bangladesh and investigate its association with important sociodemographic factors, mental health stigma and attitudes towards seeking professional psychological services. The main goal was to determine the rates of depression and anxiety measured by the PHQ-9 and GAD-7 in urban, semi-urban and rural communities. The secondary aim was to explore the relationship between sociodemographic factors, community attitudes towards mental illness (Community Attitude Toward Mental Illness—Bangla) and attitudes towards seeking professional psychological help (Attitude Toward Seeking Professional Psychological Help—Short Form) with depression and anxiety outcomes. This study was carried out to fill the evidence gap of multi-site community-based studies in Bangladesh. The study’s recruitment of adults from varied community contexts in various divisions contributes to broader, generalizable evidence needed for early identification of an individual’s risk for depression and anxiety. The results could be used to guide mental health policy and plan services in Bangladesh and to inform the development of targeted preventive mental health services. Besides, this research also offers recent evidence for future sociodemographic and regional differences in mental health of larger population groups in Bangladesh.

## 2. Materials and Methods

### 2.1 Study Design and Setting

This study was descriptive and analytical, involving a cross-section of three divisions, which were urban, semi-urban, and rural in Bangladesh. The data were collected from February 1st to June 20th, 2026, in community primary care clinics and government upazila health complexes. The three divisions chosen were Dhaka (urban), Chattogram (semi-urban), and Sylhet (rural) for the purpose of geographic comparisons of the prevalence of depression and anxiety symptoms and their sociodemographic, stigma, and help-seeking correlates.

### 2.2 Participants and Sampling

Those adults (18–60 years) attending primary care facilities were screened for eligibility. The inclusion criteria were: (1) ages 18-60 years, (2) willing to give informed consent in Bangla, and (3) absence of current psychotic episode and severe substance use disorder. A stratified multiple-stage sampling method was used where two upazilas were selected from each division purposefully, health facilities were randomly selected from each upazila, and attendees were sampled systematically until the number of participants at each health facility had reached the required sample size. For each of the facilities, the participants were stratified according to educational levels (no formal education, primary, secondary, higher secondary, bachelor’s or above) to make sure that the student population was distributed according to educational levels.

The sample size was determined by the formula n = Z^2^ p(1−p)/d^2^, with p = 0.50 (the most conservative estimate), Z = 1.96 (95% confidence level), and d = 0.05 (margin of error) resulting in a sample size of n = 384. The target was 420 (150 per division), with 10% non-response. A total of 400 participants (133–134 per division) completed the survey, with a completion rate of 95%.

### 2.3 Measurement Instruments

Depression was measured by the validated Bangla version of PHQ-9 (Chowdhury et al., 2015) and categorized as moderate or severe depression (scores ≥10) and none or mild depression (scores <10). The PHQ-9 includes nine items (rated 0-3 = not at all, 0-3 = rarely, 0-3 = a few days, 0-3 = more than a few days, 0-3 = nearly every day) and total scores ranging from 0-27.

Generalized Anxiety Disorder-7 (GAD-7; Spitzer et al., 2006) was used to measure anxiety, in which moderate to severe anxiety was considered anxiety score ≥10 and none/mild anxiety as score <10. The GAD-7 consists of seven items (all with a 4-point scale from 0 = not at all, 3 = nearly every day), with the total score ranging from 0 to 21.

The stigma of mental health was measured by 12 items of Community Attitudes Toward Mental Illness—Bangla (CAMI-B; Sarkar et al., 2018), which were answered on a 4-point Likert scale (1 = strongly disagree, 4 = strongly agree). Two items (D6 and D10) were reverse-scored prior to analysis. The total scores are in the range of 12-48, reflecting the level of stigmatizing attitudes. In this sample, the CAMI-B had a satisfactory internal consistency (Cronbach’s α = 0.695).

The Attitudes Toward Seeking Professional Psychological Help—Short Form (ATSPPH-SF; Fischer & Turner, 1970) was used to assess help-seeking attitudes. The original 10-item scale was modified to 8 items (α = 0.719) after removing two items with corrected item-total correlations <0.15 (items E2_R: r = 0.142; E10_R: r = −0.051). The 8-item version contains items scored on a 4-point scale (0 = disagree, 3 = agree), and total scores range from 0-24. Scores higher than 7 indicate more positive attitudes towards seeking professional psychological help. The internal consistency of all instruments was acceptable. Cronbach’s alpha coefficients were: PHQ-9 (α = 0.843), GAD-7 (α = 0.865), CAMI-B (α = 0.695), and ATSPPH-SF (α = 0.719). This lower coefficient for CAMI-B is due to its multidimensional nature with measurement of different aspects of stigma (Sarkar et al. 2018).

### 2.4 Data Collection

The structured questionnaire was administered by trained research assistants using face-to-face interviews in Bangla. The interview lasted about 20-25 minutes for each participant. Data gathered included sociodemographic details, which included age, sex, educational attainment, household income, employment status, marital status, religion, family type, and having access to a smartphone. All questionnaire items (PHQ-9, GAD-7, CAMI-B, and ATSPPH-SF) were administered to each participant. Paper questionnaires were used to gather data, which were then double-entered into SPSS for verification. A safety protocol was put in place: if the participant checked ≥1 on PHQ-9 item 9 (suicidal ideation), he or she was immediately clinically referred.

### 2.5 Statistical Analysis

All the sociodemographic, scale, and outcome variables were computed using descriptive statistics such as frequency, percentages, mean, and standard deviation. The continuous depression score (PHQ-9) and anxiety score (GAD-7) were transformed to binary outcome variables with moderate to severe depression and anxiety scores as the cutoff points.

Associations between the sociodemographic variables and depression/anxiety status were analyzed using chi-square tests (categorical variables) and Mann–Whitney U tests (continuous variables). Two tests were run, one with depression (moderate to severe vs. none to moderate) as the dependent variable and another with anxiety as the dependent variable. Independent variables entered simultaneously in adjusted models were age (in years), sex (female vs. male), education level (categorical), marital status (categorical), household income (categorical), and ATSPPH-SF help-seeking attitude score (continuous) and CAMI-B stigma score (continuous).

The logistic regression assumptions were also checked: the variance inflation factors (VIFs) were all less than 3, indicating no multicollinearity, and the Hosmer-Lemeshow test was used to assess the model fit. The results are reported in terms of adjusted odds ratio (aOR) and 95% confidence interval (CI) and p-value. The level of significance was defined as p < 0.05 (two-tailed). The Nagelkerke R^2^ pseudo-R^2^ coefficient was used to measure model performance. IBM SPSS Statistics, V27 (IBM Corp., 2020) was used for all analyses.

### 2.6 Ethics Approval and Informed Consent

All participants were informed in Bangla and gave written informed consent before participating in this study, which received approval from Manabik Shahajya Sangstha (MSS), Ethical Review Committee (MSS-ERC), reference no. MSS-ERC-SFP-R-35/2026. The research followed the Declaration of Helsinki and the ethical principles of research on human subjects. Personally identifiable information was not collected in the analysis dataset, and all data were coded anonymously with individual participant codes. Paper questionnaires were placed in a locked cabinet that only authorized research personnel had access to.

## 3. RESULTS

### 3.1 Sample Characteristics

The study included 400 participants from three geographical divisions (urban, semi-urban, and rural) across Bangladesh. Table 1 presents the sociodemographic characteristics of the sample. Participants ranged in age from 19 to 63 years (M = 40.22, SD = 10.14). The sample was nearly equally distributed by sex, with 133 males (33.2%) and 134 females (33.5%), while 133 participants (33.2%) did not report sex. Education was equally distributed across all five categories (n = 80 per category, 20% each), reflecting the stratified sampling design. Residence was equally stratified: 133 (33.2%) urban, 133 (33.2%) semi-urban, and 134 (33.5%) rural participants.

**Table 1.** Sociodemographic Characteristics of Respondents (N = 400)

| Variable | n | % |
| --- | --- | --- |
| Age (M ± SD) | 40.22 ± 10.14 | 19-63 |
| Sex |  |  |
| Male | 133 | 33.2% |
| Female | 134 | 33.5% |
| Residence |  |  |
| Urban | 133 | 33.2% |
| Semi-urban | 133 | 33.2% |
| Rural | 134 | 33.5% |

### 3.2 Prevalence of Depression and Anxiety

The overall prevalence of moderate-to-severe depression (PHQ-9 ≥ 10) was 39.0% (n = 156), with a mean depression score of 8.13 (SD = 5.72, range: 0–25). The overall prevalence of moderate-to-severe anxiety (GAD-7 ≥ 10) was 29.8% (n = 119), with a mean anxiety score of 7.21 (SD = 5.14, range: 0–21). Table 2 presents the prevalence stratified by sex and residence.

**Table 2.**
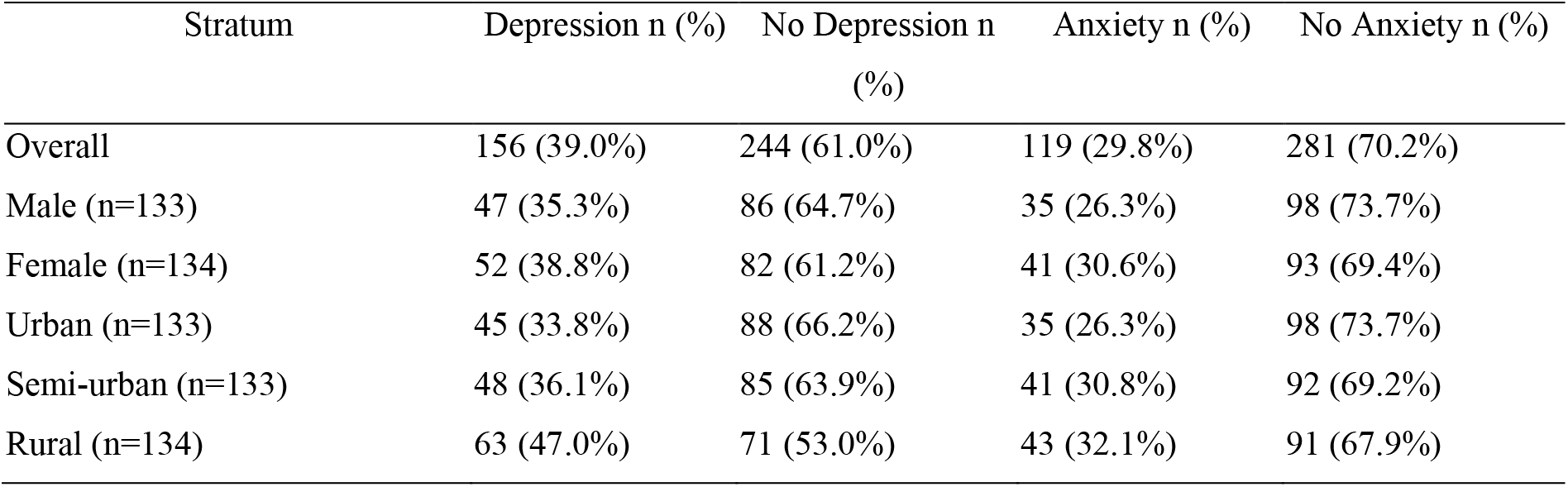
Prevalence of Depression and Anxiety by Sex and Residence (N = 400)

### 3.3 Scale Reliability

Cronbach’s alpha coefficients demonstrated acceptable internal consistency for all scales (α ≥ 0.70). PHQ-9 depression screening showed excellent reliability (α = .843), as did GAD-7 anxiety screening (α = .865). CAMI-B stigma (α = .695) and ATSPPH-SF help-seeking attitudes (α = .719) demonstrated acceptable reliability. Table 3 presents complete descriptive statistics and reliability coefficients.

**Table 3.** Reliability Coefficients and Descriptive Statistics.

| Scale | Items | $\alpha$ | M | SD | Range |
| --- | --- | --- | --- | --- | --- |
| PHQ-9 | 9 | .843 | 8.13 | 5.72 | 0-25 |
| GAD-7 | 7 | .865 | 7.21 | 5.14 | 0-21 |
| CAMI-B | 12 | .695 | 29.84 | 6.76 | 14-45 |
| ATSPPH-SF | 8 | .719 | 15.08 | 6.05 | 0-29 |

*Note.* PHQ-9 = Patient Health Questionnaire-9; GAD-7 = Generalized Anxiety Disorder-7; CAMI-B = Community Attitudes Toward Mental Illness—Bangla; ATSPPH-SF = Attitudes Toward Seeking Professional Psychological Help—Short Form.

**Table 4.**
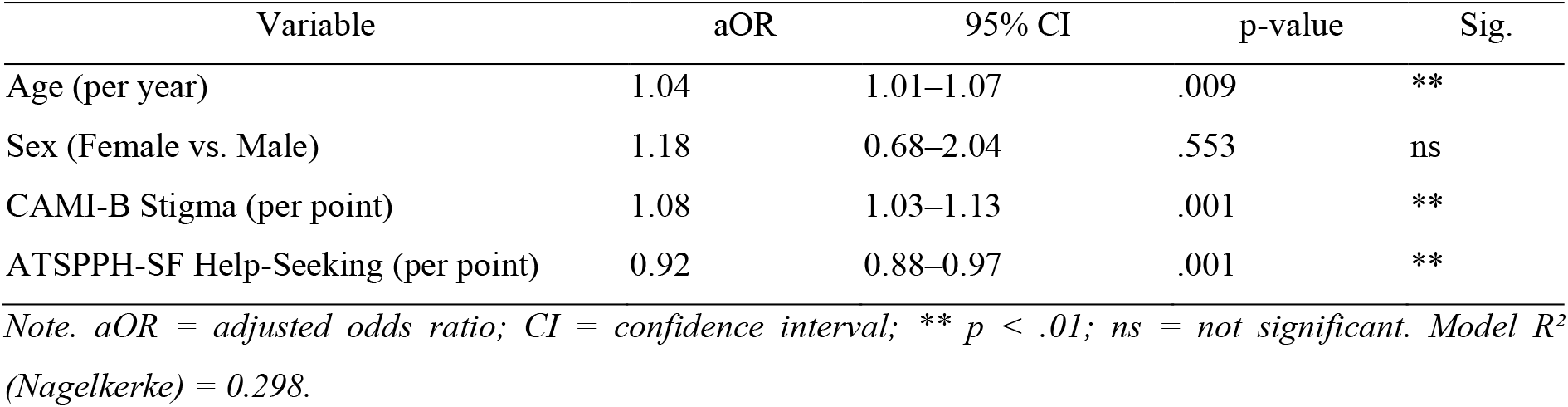
Multivariable Binary Logistic Regression: Sociodemographic Determinants of Depression (PHQ-9 ≥ 10)

**Table 5.** Multivariable Binary Logistic Regression: Sociodemographic Determinants of Anxiety (GAD-7 ≥ 10)

| Variable | aOR | 95% CI | p-value | Sig. |
| --- | --- | --- | --- | --- |
| Age (per year) | 1.02 | 0.98–1.05 | .439 | ns |
| Sex (Female vs. Male) | 1.32 | 0.73–2.37 | .352 | ns |
| CAMI-B Stigma (per point) | 1.07 | 1.02–1.12 | .004 | ** |
| ATSPPH-SF Help-Seeking (per point) | 0.93 | 0.89–0.97 | .001 | ** |

*Note.* aOR = adjusted odds ratio; CI = confidence interval; \*\* $p < .01$ ; ns = not significant. Model $R^2$ (Nagelkerke) = 0.268.

### 3.4 Logistic Regression: Determinants of Depression and Anxiety

Multivariable binary logistic regression models were fitted with depression and anxiety (dichotomized: moderate-to-severe vs. none/mild) as dependent variables. Independent variables included age, sex, education level, marital status, household income, CAMI-B stigma scores, and ATSPPH-SF help-seeking attitude scores. The models demonstrated adequate fit (Hosmer-Lemeshow test: p > .05) and acceptable multicollinearity (VIF < 3).

Multivariable models revealed that mental health stigma and help-seeking attitudes were independent predictors of depression and anxiety. Older age significantly increased odds of depression (aOR = 1.04, p = .009). Stigma increased odds of both depression (aOR = 1.08, p = .001) and anxiety (aOR = 1.07, p = .004), while positive help-seeking attitudes reduced odds of depression (aOR = 0.92, p = .001) and anxiety (aOR = 0.93, p = .001). Sex, education, and income were not significant predictors.

## 4. Discussion

The aim of this multi-site cross sectional study was to investigate the prevalence of depression and anxiety and its association with sociodemographic factors, mental health stigma and the attitudes of adults towards professional psychological help in Bangladesh. Sixty percent of the participants had moderate-to-severe depression (39.0%) and 30% had moderate-to-severe anxiety (29.8%). These results reflect a significant level of psychological symptoms among the study sample, and fall at the upper limit of regional and global prevalence estimates.

When compared with already published prevalence data in Bangladesh, important patterns can be observed. National surveys estimate the prevalence of depression at 5.2%–6.7% and anxiety at 3.5%–4.7% (Sarkar et al., 2026; Pengpid et al., 2025), compared to screening-based studies which estimate 16.3% and 6.0%, respectively (Sarkar et al., 2026). The estimates of the present study (39.0% depression, 29.8% anxiety) are significantly higher than both diagnostic and previous screening estimates. This difference is due to a number of methodological factors: (1) screening instruments (PHQ-9, GAD-7) measure dimensional symptom severity; these measures are more sensitive and include a wider range of severity than just severe cases; (2) the present sample included patients attending primary care who may have a higher symptom burden than the general population; and (3) the study criterion for PHQ-9/GAD-7 ≥10 is for moderate-to-severe symptoms, which includes a broader range of symptoms than severe symptoms alone. Specifically, the prevalence rate was highly comparable to the prevalence rates estimated from screening measures around the world: a meta-analysis of 218 studies showed pooled depression prevalence rate of 36.7% (95% CI [33.8–39.6]) using PHQ-9, and pooled anxiety prevalence rate of 30.6% (95% CI [28.5–32.8]) using GAD-7 (Levis et al., 2019; Dear et al., 2020, respectively). This alignment with the international standards validates the current findings and implies that the prevalence of the current study is similar to that in other LMIC countries based on screening.

They should be understood as prevalence estimates with regard to the design of the study and the way it was measured. Depression and anxiety were diagnosed with validated screening scales, not with structured clinical interviews. These estimates are for participants who have moderate to severe depressive or anxiety symptoms, and are not necessarily a representation of people who have a psychiatric diagnosis. Due to the cross-sectional nature of the design, conclusions on causality and time direction are not possible and the relationships between stigma, help-seeking attitudes, and psychological symptoms should be understood as associations.

Descriptive findings showed variation across demographic subgroups. Depression was reported by 35.3% of males, 38.8% of females, and 47.0% of rural participants, whilst anxiety was reported by 26.3%, 30.6%, and 32.1%, respectively. The rural-urban gradient observed for depression (47.0% rural vs. 33.8% urban vs. 36.1% semi-urban) merits particular attention. Prior Bangladeshi research has documented rural-urban differences in mental health prevalence, though findings have been mixed (Islam, 2019; Amin et al., 2025). A nationally representative analysis by Amin et al. (2025) found no consistent rural-urban gradient in depression and anxiety prevalence after controlling for sociodemographic characteristics. The present study similarly found that whilst rural residence showed elevated descriptive prevalence, it did not emerge as a significant independent predictor in multivariable models. This suggests that the observed rural-urban disparity is partially confounded by or mediated through stigma, help-seeking attitudes, or other unmeasured psychosocial factors, rather than representing a direct environmental or geographic effect.

Interestingly, there was no significant relationship between depression or anxiety and sex in the adjusted models. This is an unprecedented discovery in the extensive literature in Bangladesh and South Asia. In university and school-based samples, women were found to have higher levels of depression and anxiety (Islam et al., 2020, 2021) and a number of population-based studies reported female predominance in depression (Amin et al., 2025; Pengpid et al., 2025). Present findings, however, are consistent with certain studies indicating that after adjusting for psychosocial factors, sex differences in prevalence of depression/anxiety are reduced or eliminated. A key hypothesis to be raised from this lack of sex effect after controlling for stigma and attitudes about help-seeking is that there may be a portion of the sex difference in many studies that is due to differential exposure to stigma and help-seeking attitudes by sex, but is not a sex effect. However, under conditions of equal stigma and attitudes, the sex effect is reduced; in Bangladesh, cultural gender norms may lead women to have higher levels of perceived stigma and more negative help-seeking attitudes, which could result in higher reporting of symptoms and more help-seeking. This interpretation implies that gender-specific stigma reduction and/or help-seeking enhancement interventions could help close apparent sex disparities in mental health outcomes.

Age emerged as a significant and independent predictor of depression but not anxiety. Each additional year of age was associated with a 4% increase in the odds of depression (aOR = 1.04, 95% CI [1.01–1.07], p = .009), whilst age was not significantly associated with anxiety (aOR = 1.02, 95% CI [0.98–1.05], p = .439). This age-depression association is consistent with global epidemiological trends (Jain et al., 2022; Chodavadia et al., 2023) and Bangladesh-specific research. Amin et al. (2025) found a linear age-depression relationship among women in nationally representative data, and Islam (2019) documented higher depressive symptoms in older compared to younger adults in rural Bangladesh. The lack of age effect on anxiety is noteworthy and less commonly reported. Possible explanations include: (1) anxiety may be more uniformly distributed across the lifespan; (2) different anxiety subtypes (generalized anxiety, social anxiety, panic) may show different age trajectories; (3) cohort effects or survival bias (individuals with severe anxiety may have different mortality or healthcare-seeking patterns). This divergence between depression and anxiety age effects warrants further investigation in future research.

Education and household income were not significant independent predictors of depression or anxiety in adjusted analyses. Whilst socioeconomic disadvantage has been consistently associated with poor mental health in literature from high-income countries (Jain et al., 2022) and some low- and middle-income settings (Arvind et al., 2019), the relationship is less consistent in Bangladesh and South Asia. In the present study, education and income showed no association with depression or anxiety after controlling for stigma and help-seeking attitudes. Similarly, Amin et al. (2025) found that among women, education and income were not independently associated with depression and anxiety prevalence in nationally representative data. This pattern contrasts with findings in India, where Arvind et al. (2019) found strong associations between low education, low income, and depression prevalence in multisite data. The lack of socioeconomic effect in Bangladesh may reflect: (1) mental health services being largely unaffordable and inaccessible regardless of income level, such that economic barriers to care do not translate into symptom differences; (2) strong role of cultural factors (stigma, attitudes, beliefs) that cut across socioeconomic strata; (3) measurement or sample composition factors. This finding underscores that in resource-limited contexts, conventional sociodemographic characteristics alone may inadequately explain mental health variation, and psychosocial factors warrant greater emphasis.

The most consistent and striking associations were observed for mental health stigma and help-seeking attitudes— findings that represent a key novelty and strength of this research. Higher CAMI-B stigma scores were independently associated with both depression and anxiety. Each one-point increase in stigma was associated with an 8% increase in the odds of depression (aOR = 1.08, 95% CI [1.03–1.13], p = .001) and a 7% increase in the odds of anxiety (aOR = 1.07, 95% CI [1.02–1.12], p = .004). Conversely, higher ATSPPH-SF scores (indicating more positive help-seeking attitudes) were associated with lower odds of both depression and anxiety. Each one-point increase in help-seeking attitude score was associated with an 8% reduction in the odds of depression (aOR = 0.92, 95% CI [0.88–0.97], p = .001) and a 7% reduction in the odds of anxiety (aOR = 0.93, 95% CI [0.89– 0.97], p = .001). These parallel and opposing associations across two psychosocial domains are particularly noteworthy: they demonstrate that stigma and help-seeking attitudes are independent psychological correlates of depression and anxiety in this population.

The stigma findings align with and extend prior Bangladesh research. Faruk et al. (2023) documented substantial mental illness stigma in Bangladesh, finding that discriminatory attitudes, perceptions of dangerousness, and beliefs about incurability are common. Roy & Chowdhury (2024) similarly reported widespread stigmatizing attitudes in the general population. The present study advances this literature by demonstrating a quantitative association between community-level stigma attitudes and individual-level depression and anxiety symptoms. Cross-culturally, research has established that stigma correlates with psychological distress (Alluhaibi & Awadalla, 2022), and in South Asian contexts, stigma toward mental illness is a recognized barrier to treatment engagement (Vidyasagaran et al., 2023). The magnitude of association in the present study (aOR ≈ 1.07–1.08 per scale point) is substantial from a public health perspective: a person with stigma scores at the 75th percentile versus the 25th percentile would have approximately double the odds of depression or anxiety, representing a clinically meaningful effect size.

The help-seeking attitude findings are particularly important and, to the authors’ knowledge, represent one of the first multi-site quantitative documentations of this association in Bangladesh. Prior qualitative research has identified help-seeking barriers related to stigma, low mental health awareness, and perceptions that professional care is ineffective (Ahmed et al., 2025; Faruk et al., 2023). The present study operationalizes and quantifies these attitudes using a validated instrument and demonstrates that positive help-seeking attitudes are protective against depression and anxiety. This protective effect aligns with Social Cognitive Theory, suggesting that individuals who view professional help as appropriate, efficacious, and non-stigmatizing are better equipped psychologically to manage distress. Notably, these associations remained independent and statistically significant even after controlling for sociodemographic factors, highlighting the importance of psychological and attitudinal variables in explaining mental health variation in Bangladesh.

### Theoretical Frameworks

The findings are consistent with the established theories from psychology and public health that explain the mental health in low and middle income contexts. A helpful concept that frames the ideas is the Stress and Coping framework (Lazarus & Folkman, 1984). In this context, mental health stigma acts as a chronic social stressor, which increases psychological vulnerability. If an individual internalizes stigmatizing messages, whether through the community, through the family, through personal statements, they’ll feel shame, social isolation, positive expectations about discrimination, and a decreased desire to disclose symptoms or reach out for help. The following are well documented psychological processes that intensify and sustain depression and anxiety symptoms (Corrigan & Penn, 1999): The avoidance and internalized shame that might stem from cultural beliefs about mental illness as stemming from moral weakness, family shame, supernatural, etc. or social karma, is especially relevant in the context of Bangladesh where mental illness is often explained in terms of moral failure (Faruk et al., 2023; Roy & Chowdhury, 2024). The current study results supporting this theoretical mechanism are empirical as they suggest that stigma is an independent predictor of depression and anxiety.

Positive attitudes towards help-seeking is a protective factor that is consistent with the Social Cognitive Theory (Bandura, 1986) and the concept of self-efficacy. In this understanding, people who have high self-efficacy in coping with health issues show greater coping and emotional regulation, specifically because they feel confident in their ability to access professional help and medical services when needed, and that it would be effective and socially accepted. Self-efficacy beliefs are implicated in whether or not individuals: (1) seek help when upset; (2) continue to utilize treatment; and (3) sustain therapeutic benefits over time. In Bangladesh’s mental health service environment where there is limited mental health services and high stigma, the individuals with positive help seeking attitudes may be fundamentally different in their psychological appraisals of distress, instead of internalizing self blame and internalized shame (depression/anxiety) they might reframe the symptoms as manageable medical problems that are amenable to treatment. This cognitive reframing and consequent agency may offer a psychological barrier to symptom escalation. On the other hand, those with negative help-seeking attitudes are stuck in a psychological rut – they suffer silently, don’t seek professional help, and thus suffer from long and possibly worsening symptoms, as found in qualitative studies conducted in Bangladesh (Ahmed et al., 2025).

Developmental and life-course perspectives may be used to explain the age depression finding not the anxiety finding. In the older age group (mid-to-older adulthood) in Bangladesh, a combination of psychosocial stress factors emerge: marital disruption (divorce, separation), bereavement (loss of spouse and/or children), decreased roles and purpose in society, health decline and chronic illness, and economic insecurity (pension insufficient or loss of employment status). These experiences are directly and consistently identified as a part of depressive symptomology in various cultures (Amin et al., 2025; Islam, 2019). By comparison, anxiety can be fairly consistent across the life span or can have different age patterns, depending on the type of anxiety and life circumstances. After accounting for the sex effect by stigma and attitudes, the lack of this effect indicates that observed sex differences in depression and anxiety prevalence in many studies might be partly through a mediation pathway of sex differences in stigma and attitudes to help-seeking, rather than sex being a primary or independent risk factor. This has profound implications for intervention, as interventions to reduce stigma and attitudes for mental health could go a long way towards reducing disparities by targeting underlying psychological mechanisms or processes.

### Limitations

There are four important restrictions that should be noted. Firstly, the cross-sectional design cannot make causal inferences. But it could be the other way around; depression and anxiety can be the causes of perceived stigma or of the attitudes of help seeking, not stigma and attitudes the causes of depression and anxiety. The observed correlations may also be due to unmeasured confounding (e.g., another factor may be associated with stigma and mental illness at the same time). To determine direction of effect and causality requires longitudinal studies with temporal sequencing.

Secondly, the sample was collected from the primary care consumers of government health care facilities, which creates selection bias. There is likely some variation in health awareness, symptom burden, and the likelihood of seeking health care for health care seekers when compared to the general population. Thus, the results of the findings cannot be generalised to the larger Bangladeshi adult population, especially the non-formal health care access population. More generalizability could be provided through community-based household sampling designs.

Third, depression and anxiety were measured by screening measures (PHQ-9, GAD-7), which may have led to an overestimate of prevalence. However, screening instruments are suitable for use in estimating prevalence in a population, comply with the WHO recommendations and allow for comparison between studies.

Fourth, the stratified sampling by education ensured equal representation across categories (n = 80 per category, 20% each), which does not reflect the population’s true education distribution and may limit generalizability of education-based prevalence estimates to the broader population.

### Implications for Intervention and Policy

The implications of this research for intervention and policy will be discussed. The high rates of depression (39.0%) and anxiety (29.8%) along with the respective independent predictive ability of stigma and help-seeking attitudes highlight the need to draw urgent attention to comprehensive, multi-level mental health interventions in Bangladesh. Based on the present results, there are a few directions for evidence-based interventions.

Community-level stigma reduction: The public mental health campaigns should engage with and counteract cultural beliefs regarding causes of mental illness, dangerousness and treatability. Contact, education, protest (Corrigan & Penn, 1999) are stigma reduction strategies that should be culturally adapted for Bangladesh. Campaigns should focus on the fact that mental illness is treatable, common, and it is not a personal or family disgrace. Religious leaders, community elders, and respected community members as messengers can increase cultural legitimacy and reach. Existing literature from South Asia indicates that contact-based interventions which utilize personal recovery stories of those with lived mental health experiences have a significant impact on stigma (Vidyasagaran et al., 2023).

Individual Help-Seeking Enhancement: Include explicit targeting of help seeking attitudes in psychological interventions using psychoeducation, motivational interviewing, and peer support. Reducing access barriers through low-intensity digital interventions (phone-based counselling, culturally adapted, mental health apps). The inclusion of trusted persons such as family members, religious counselors and female health workers can help improve acceptability, especially among women where cultural barriers to help-seeking exist. Brief DMLPs shown to be feasible and acceptable in Bangladesh (Habib et al., 2025).

Healthcare System Strengthening: The multi-site design documented geographic prevalence variation (rural >semi-urban>urban for depression), highlighting need for expanded mental health services, particularly in rural areas. Task-sharing models—training non-specialist health workers to deliver evidence-based treatments—are cost-effective and scalable. Integration of PHQ-9/GAD-7 screening into routine primary care facilitates early identification and linkage to care. Brief psychological interventions in primary care settings reduce the need for specialist referral and improve accessibility.

Age and Gender-Specific Strategies: Age-specific stressors (e.g., bereavement, health decline, economic insecurity) warrant interventions for older adults who demonstrate a high risk for depression. The involvement of families and peer support can increase engagement. Changing the approach to using community health worker outreach to women’s groups or trusted female mentors may help boost engagement among women with specific help-seeking barriers.

## 5. Conclusion

This multi-site cross-sectional study revealed a significant prevalence of depression (39.0%) and anxiety (29.8%) among the adults in urban, semi-urban, and rural areas of Bangladesh. The rates of depression and anxiety are consistent with international screening-derived rates, reinforcing the reliability of results and comparability with other countries’ epidemiological data. Age was a predictor of depression, but not anxiety; age alone was not an independent predictor in models that controlled for other conventional sociodemographic characteristics (sex, education, income). Most importantly, mental health stigma and attitudes towards professional psychological assistance were found to be consistent, independent, and psychologically meaningful factors influencing depression and anxiety risk levels. The reduction of mental health burden in Bangladesh should be achieved through the integration of culturally appropriate and comprehensive psychosocial interventions at the community level, focused on reducing stigma, and individual-based interventions focused on improving help-seeking attitudes, as well as through strengthening of health care systems and digital innovation. Concomitant with increasing service availability and accessibility, by tackling these modifiable psychosocial risk factors, Bangladesh can aim to reduce the significant treatment gap for mental health and enhance mental health outcomes for adults.

## Data Availability

All data produced in the present study are available upon reasonable request to the authors

## Acknowledgments

We acknowledge the support of government upazila health complexes and community primary care clinics in Dhaka, Chattogram, and Sylhet divisions for facilitating data collection. We thank the research assistants who conducted participant interviews and data entry. We acknowledge the Bangladesh Medical Research Council for ethical oversight.

## Declaration of AI Use

During the preparation of this work, the authors used Claude (Anthropic), an artificial intelligence language model, to support writing and as an organizational assistant. All scientific decisions, data analyses, interpretations, and conclusions remain the responsibility of the human authors. The authors have critically reviewed all AI-assisted text and take full responsibility for the accuracy and integrity of the manuscript.

## Author Contributions

Sabbir Maruf: Conceptualization, Study Design, Methodology, Data Collection, Supervision, Data Analysis, Writing - Original Draft, Writing - Review & Editing. Habib: Conceptualization, Methodology, Writing - Review & Editing. Faria: Data Collection, Introduction and Conclusion. Shakila: Results and Discussion. Mamunur: Abstract and Conclusion; also Masum and Bayezid: Data Collection

## Data Availability Statement

The datasets generated during the current study are not publicly available due to participant confidentiality and ethical restrictions but are available from the corresponding author on reasonable request with appropriate institutional ethics approval and data sharing agreements.

